# SEX DIFFERENCES IN PHARMACOLOGIC OPTIMAL MEDICAL THERAPY FOR ISCHEMIC HEART DISEASE, 2010–2020: AN OBSERVATIONAL STUDY

**DOI:** 10.1101/2025.07.08.25331126

**Authors:** Hassan A. Alhassan, Harnoor Mann, Leonard Chiu, Bary Malik, Malamo Countouris, Amber E Johnson

## Abstract

**Background:** Despite guideline recommendations for optimal medical therapy (OMT) in the secondary prevention of ischemic heart disease (IHD)—including antiplatelets, statins, renin-angiotensin-aldosterone system inhibitors (RAASi), and β-blockers—substantial sex disparities in OMT utilization persist. The extent to which national efforts have mitigated these disparities in contemporary cohorts remains unclear.

**Methods:** We analyzed data from the 2011–2020 National Health and Nutrition Examination Survey (NHANES) cycles, identifying adults with self-reported IHD (defined as a history of myocardial infarction or coronary heart disease). OMT use in the preceding 30 days was assessed based on participant report and verified through medication containers when available. We evaluated trends in individual drug classes and common combinations, stratified by sex.

**Results:** Among 1,905 adults (mean age 65.4 years; 40.6% women), women had significantly lower rates of OMT use compared to men, including antiplatelets (68.0% vs 77.7%), statins (57.2% vs 73.9%), RAASi (45.6% vs 59.0%), and β-blockers (51.2% vs 61.1%). Women were also less likely to use guideline-recommended combinations such as aspirin plus statins (47.4% vs 64.1%) and all four OMT classes (17.5% vs 32.5%). After adjustment for sociodemographic and clinical factors, women remained less likely to use antiplatelets (OR 0.71; 95% CI, 0.52– 0.94), statins (OR 0.62; 95% CI, 0.40–0.96), and RAASi (OR 0.56; 95% CI, 0.38–0.84), while β-blocker use did not differ significantly. These sex-based disparities were consistent across all survey cycles from 2011 to 2020.

**Conclusion:** In this nationally representative study, women with IHD were significantly less likely than men to receive guideline-directed OMT, with persistent disparities over the past decade. These findings underscore the need for targeted strategies to close the sex gap in cardiovascular prevention.

## Introduction

Ischemic heart disease (IHD) remains the leading cause of cardiovascular mortality and morbidity in the United States (US), accounting for over 380,000 deaths in 2020.^1^ While IHD mortality rates in the US and globally are lower among women than men, women consistently experience higher rates of recurrent major adverse cardiovascular events. A significant contributing factor to this sex difference in outcomes is the suboptimal use of optimal medical therapy (OMT), including antiplatelet agents, lipid-lowering medications such as HMG-CoA reductase inhibitors (statins), beta-blockers (β-blockers), and renin-angiotensin-aldosterone system inhibitors (RAASi).^2,3–6^.Moreover, recent studies show that young women (≤50 years) with obstructive coronary disease have a higher rate of complications and mortality compared to young men.^7–9^ These observations highlight a dire need to understand management patterns in this cohort as a means of reducing not only the disease burden but also the economic implications on their families and the health care system.

Over the past few decades, there have been numerous national initiatives aimed at addressing sex disparities in the care of cardiovascular diseases (CVD). The Go Red for Women campaign was launched in 2004 by the American Heart Association (AHA) to raise awareness of heart disease in women.^10^ In that same year, the AHA issued the first evidence-based guidelines for preventing CVD in women.^11^ This was followed by an effectiveness-based guidelines update in 2011, which emphasized the need for aggressive and equitable management including the need to reach targeted doses of evidence-based medications among women.^12^

However, the extent to which these national efforts have effectively closed the sex gap in OMT use remains unclear. Beyond awareness, the impact of major health policy initiatives, such as the Affordable Care Act, on insurance and medication coverage, and their role in addressing this disparity, is also not well established.

While previous studies have identified persistent sex differences in the use of some OMT, few studies have examined trends in the use of OMT combinations in a contemporary national database.^13–15^ We sought to comprehensively address this gap in the literature by pursuing three objectives: (1) to assess sex differences in the use of individual OMT classes and guideline-recommended combinations; (2) to evaluate sex-specific trends in OMT use from 2011 to 2020; and (3) to compare the prevalence of OMT use by sex across age groups (<50 years and ≥50 years).

## Methods

We performed a serial cross-sectional analysis using aggregate data from the National Health And Nutrition Examination Survey (NHANES) cycles from 2011 through 2020. Briefly, NHANES is one of the nationally representative National Center for Health Statistics data surveys, conducted serially on a sample of non-institutionalized civilian adults in the US. A detailed description of the NHANES protocols and methodology can be found at https://www.cdc.gov/nchs/nhanes/index.htm. Because the data are publicly available and deidentified, the Institutional Review Board at the University of Pittsburgh determined that the study was exempt from review. We followed the Strengthening the Reporting of Observational Studies in Epidemiology checklist.^16^ Our analytic codes are available upon reasonable request.

### Study cohort

We included all non-pregnant adults (aged ≥18 years) with IHD, defined as a self-reported history of myocardial infarction or coronary heart disease at the time of survey administration. Pregnant women were excluded because some of the medications studied are contraindicated in pregnancy. Details of the study cohort creation are summarized in **Figure 1**.

**Figure 1:**
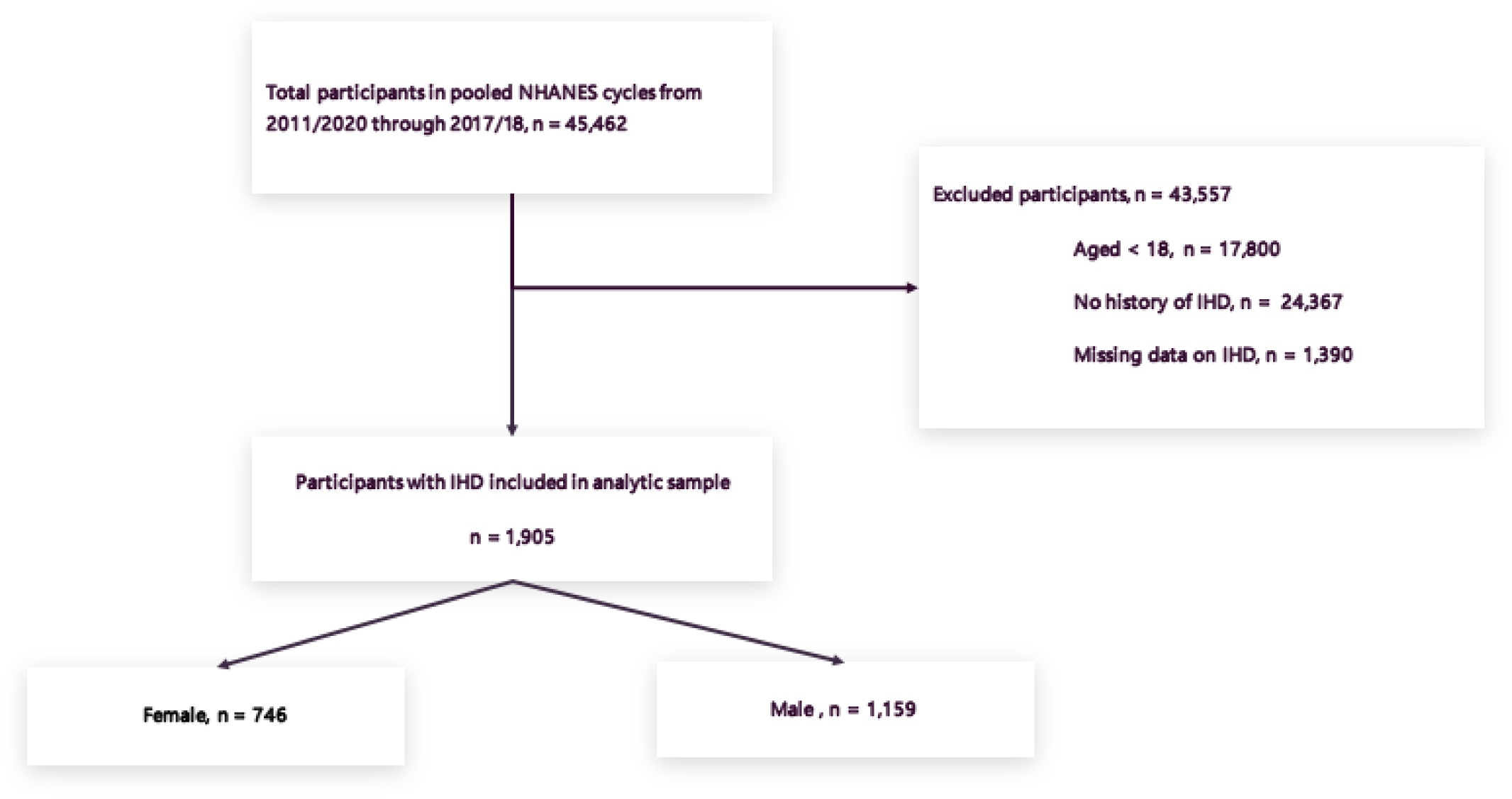
Flow chart summarizing study cohort creation.

### Outcome

The protocol for medication review in the NHANES has been described previously.^17^ Briefly, data on prescription medications used in the past 30 days were collected during household interviews. Participants who reported using medication were asked to show the medication containers to the interviewer when available. All medications were converted to generic names and categorized into standard therapeutic classes using the Multum MediSource Lexicon classification system. Combination drugs were included in their component classes, consistent with prior studies.^18^ We used therapeutic codes to classify OMT, defined as use of antiplatelets, statins, RAASi, and β-blockers, following AHA/ACA guidelines for secondary ASCVD prevention. Antiplatelets included self-reported and verified use of aspirin, clopidogrel, prasugrel, or ticagrelor. RAASi included angiotensin receptor blockers and angiotensin-converting enzyme inhibitors. β-blockers included cardioselective agents such as carvedilol, bisoprolol, and metoprolol.

### Exposure and covariables

The NHANES collects data on gender as self-reported ‘male’, ‘female’, or missing, without clearly distinguishing between sex assigned at birth and gender. Given that subsequent NHANES questionnaire items referred to sex assigned at birth rather than gender, we used the sex variable when available.

NHANES data also include age, self-reported race and ethnicity (non-Hispanic white, non-Hispanic Black, Mexican American, Asian American, or other), level of educational attainment (high school or less, some college, or college graduate), and annual family income (which was relative to the federal poverty level (FPL) and adjusted for family size and survey year. Income was classified as: <130% of FPL, 130-349% of FPL, or ≥350% of the FPL). Additional covariates were health insurance status, and current smoking status. We defined clinical variables as follows: history of hypertension was determined by self-reported use of antihypertensive medication, a self-reported diagnosis of hypertension, or an average of three blood pressure measurements of ≥130/80 mm Hg; diabetes as self-reported use of antidiabetic medication, a self-reported diagnosis of diabetes, or a hemoglobin A1c level of ≥6.5%; hyperlipidemia as low-density lipoprotein cholesterol ≥160 mg/dL or non–high-density lipoprotein cholesterol ≥ 190 mg/dL; obesity was defined as body mass index (BMI) ≥ 30 kg/m^2^; depression as an aggregate Patient Health Questionnaire (PHQ) 9 score of ≥10; chronic kidney disease (CKD) as an estimated glomerular filtration rate of ≤ 60 mL/min/1.73m^2^ (calculated using the CKD-EPI equation^19^).^20^

### Statistical analysis

We compared sociodemographic and health characteristics across sex using weighted linear regression models. We presented continuous variables as weighted means and standard errors, and categorical variables as weighted percentages and standard errors. We compared baseline characteristics across sex using simple weighted linear or logistic regression models.

We then calculated sex-specific prevalence estimates of OMT use separately and in aggregate using simple regression models. To estimate the odds ratios for each class of OMT, we used weighted multiple logistic regression models, adjusting for demographics (age, race/ethnicity), comorbidities (hypertension, diabetes, obesity, hyperlipidemia, smoking status, depression, and CKD), socioeconomic status (level of education attained, annual family income, and health insurance status), and survey-period to account for temporal trends. To estimate odds ratios for each class of OMT, we used weighted multivariable logistic regression models adjusting for demographics (age, race/ethnicity), comorbidities (hypertension, diabetes, obesity, hyperlipidemia, smoking status, depression, chronic kidney disease), socioeconomic factors (education, income, insurance status), and survey period to account for temporal trends. These covariates were included to isolate the independent association between sex and OMT use. To test for differences in OMT use over time by sex, we included a multiplicative term for sex and each survey period (modeled as the midpoint of each survey period) in a regression model with medication use as the outcome variable.

To assess non-linear trends, we included a quadratic term for survey year. In subgroup analysis, we examined sex-differences in prevalence and trends of OMT use by repeating the above analysis stratified by age categories: <50 years and ≥50 years.

Given the low proportion of missing data—hypertension (7.9%), obesity (7.4%), chronic kidney disease (8.5%), education (<1%), and health insurance status (<1%)—and the absence of missing values for other covariates, we used a complete-case analysis approach to address missingness in the data.

In all analyses, we accounted for the complex sampling design of NHANES and used Taylor series linearization to calculate confidence intervals. We used the survey specific procedures in STATA version 17

(StataCorp, College Station, TX), with statistical significance defined as a two-sided α = 0.05.

## Results

The response rate for interviewed participants ranged from 51.0%-72.6% and with medication containers verified in 82.3%-89.4% of cases. Of the 1,905 adults with IHD (representing 14.2 million US adults) included in the final analytic sample, 40.6% were women, the mean age was 65.4 years, 74.5% self-identified as non-Hispanic White adults, and 53.4% were covered by Medicare. Women were more likely to identify as non-Hispanic Black adults, carry comorbid depression, CKD, or hyperlipidemia, have Medicaid as their primary health insurer, or report an annual income level <130% of FPL. **Table 1** summarizes the characteristics of the study participants.

**Table 1:**
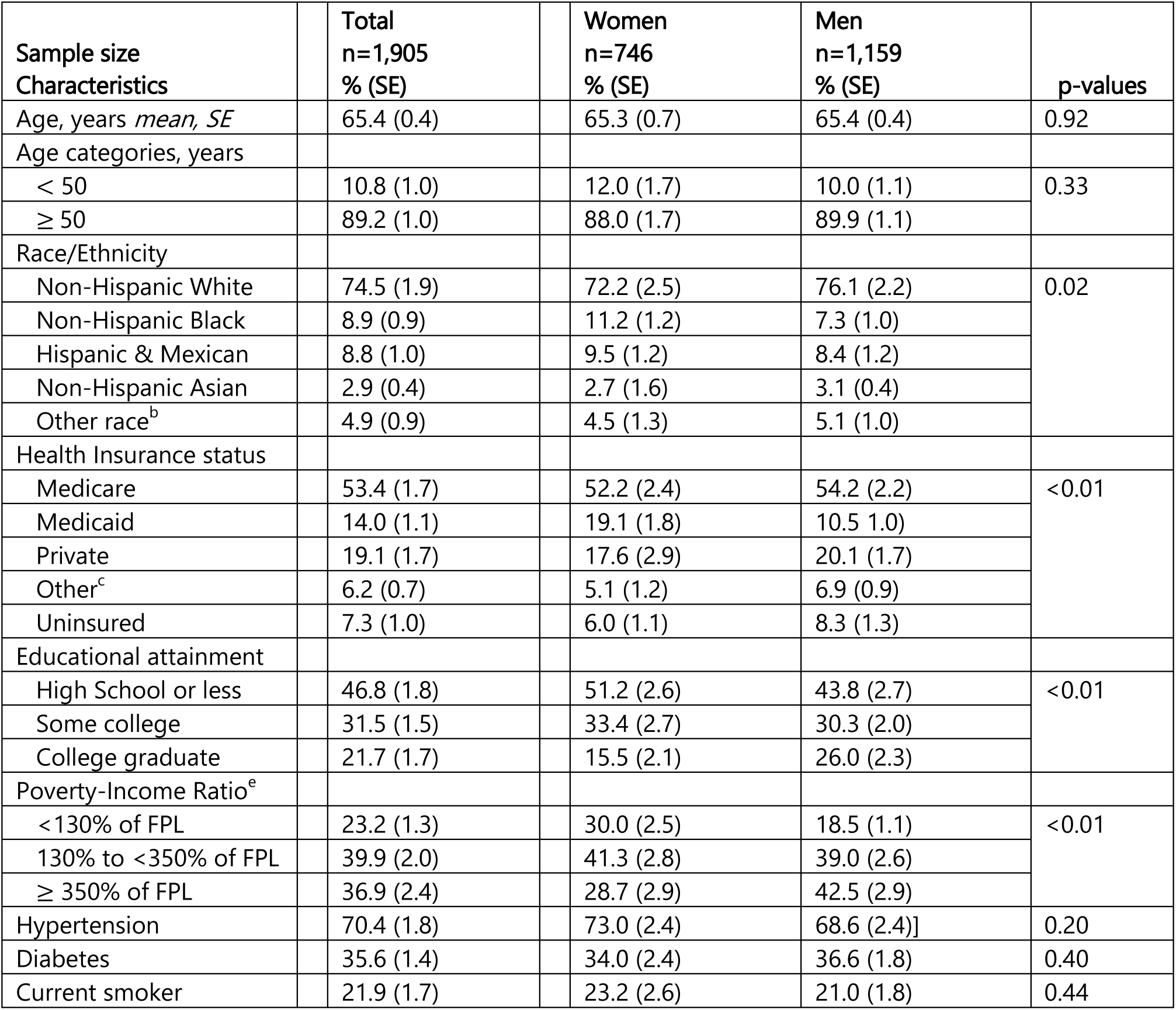
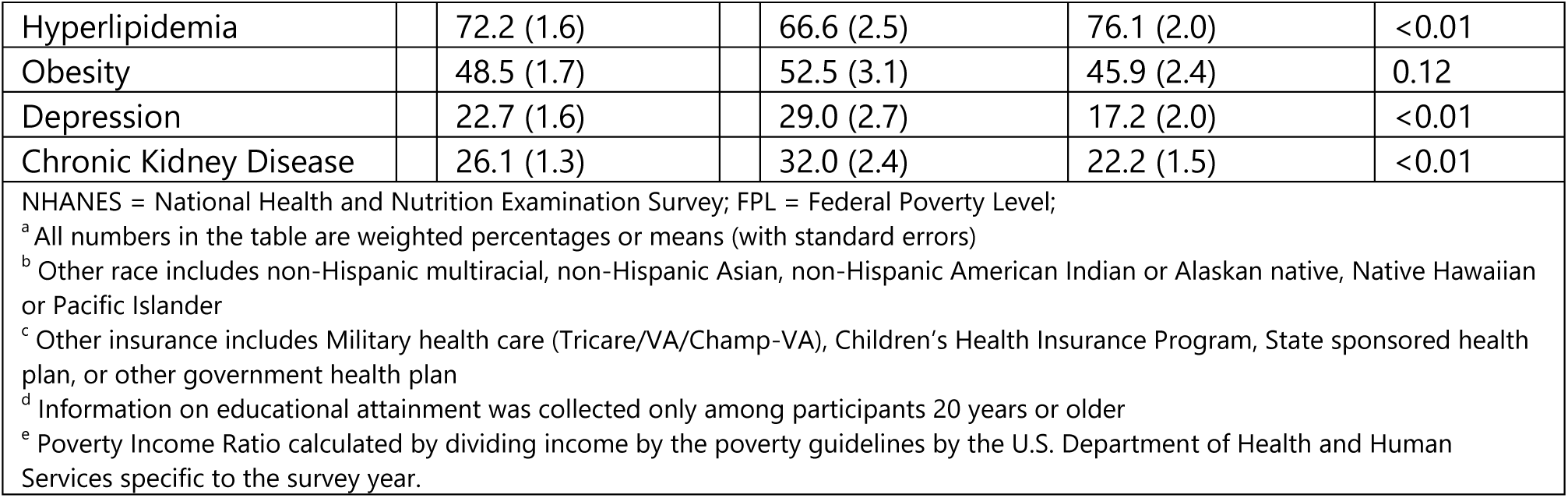
Study participants’ characteristics.

| Sample size<br>Characteristics | Total<br>n=1,905<br>% (SE) | Women<br>n=746<br>% (SE) | Men<br>n=1,159<br>% (SE) | p-values |
| --- | --- | --- | --- | --- |
| Age, years *mean, SE* | 65.4 (0.4) | 65.3 (0.7) | 65.4 (0.4) | 0.92 |
| Age categories, years |  |  |  |  |
| < 50 | 10.8 (1.0) | 12.0 (1.7) | 10.0 (1.1) | 0.33 |
| ≥ 50 | 89.2 (1.0) | 88.0 (1.7) | 89.9 (1.1) |  |
| Race/Ethnicity |  |  |  |  |
| Non-Hispanic White | 74.5 (1.9) | 72.2 (2.5) | 76.1 (2.2) | 0.02 |
| Non-Hispanic Black | 8.9 (0.9) | 11.2 (1.2) | 7.3 (1.0) |  |
| Hispanic & Mexican | 8.8 (1.0) | 9.5 (1.2) | 8.4 (1.2) |  |
| Non-Hispanic Asian | 2.9 (0.4) | 2.7 (1.6) | 3.1 (0.4) |  |
| Other race<sup>b</sup> | 4.9 (0.9) | 4.5 (1.3) | 5.1 (1.0) |  |
| Health Insurance status |  |  |  |  |
| Medicare | 53.4 (1.7) | 52.2 (2.4) | 54.2 (2.2) | <0.01 |
| Medicaid | 14.0 (1.1) | 19.1 (1.8) | 10.5 (1.0) |  |
| Private | 19.1 (1.7) | 17.6 (2.9) | 20.1 (1.7) |  |
| Other<sup>c</sup> | 6.2 (0.7) | 5.1 (1.2) | 6.9 (0.9) |  |
| Uninsured | 7.3 (1.0) | 6.0 (1.1) | 8.3 (1.3) |  |
| Educational attainment |  |  |  |  |
| High School or less | 46.8 (1.8) | 51.2 (2.6) | 43.8 (2.7) | <0.01 |
| Some college | 31.5 (1.5) | 33.4 (2.7) | 30.3 (2.0) |  |
| College graduate | 21.7 (1.7) | 15.5 (2.1) | 26.0 (2.3) |  |
| Poverty-Income Ratio<sup>e</sup> |  |  |  |  |
| <130% of FPL | 23.2 (1.3) | 30.0 (2.5) | 18.5 (1.1) | <0.01 |
| 130% to <350% of FPL | 39.9 (2.0) | 41.3 (2.8) | 39.0 (2.6) |  |
| ≥ 350% of FPL | 36.9 (2.4) | 28.7 (2.9) | 42.5 (2.9) |  |
| Hypertension | 70.4 (1.8) | 73.0 (2.4) | 68.6 (2.4) | 0.20 |
| Diabetes | 35.6 (1.4) | 34.0 (2.4) | 36.6 (1.8) | 0.40 |
| Current smoker | 21.9 (1.7) | 23.2 (2.6) | 21.0 (1.8) | 0.44 |

|  |  |  |  |  |  |
| --- | --- | --- | --- | --- | --- |
| Hyperlipidemia |  | 72.2 (1.6) | 66.6 (2.5) | 76.1 (2.0) | <0.01 |
| Obesity |  | 48.5 (1.7) | 52.5 (3.1) | 45.9 (2.4) | 0.12 |
| Depression |  | 22.7 (1.6) | 29.0 (2.7) | 17.2 (2.0) | <0.01 |
| Chronic Kidney Disease |  | 26.1 (1.3) | 32.0 (2.4) | 22.2 (1.5) | <0.01 |
| NHANES = National Health and Nutrition Examination Survey; FPL = Federal Poverty Level;<br><sup>a</sup> All numbers in the table are weighted percentages or means (with standard errors)<br><sup>b</sup> Other race includes non-Hispanic multiracial, non-Hispanic Asian, non-Hispanic American Indian or Alaskan native, Native Hawaiian or Pacific Islander<br><sup>c</sup> Other insurance includes Military health care (Tricare/VA/Champ-VA), Children's Health Insurance Program, State sponsored health plan, or other government health plan<br><sup>d</sup> Information on educational attainment was collected only among participants 20 years or older<br><sup>e</sup> Poverty Income Ratio calculated by dividing income by the poverty guidelines by the U.S. Department of Health and Human Services specific to the survey year. |  |  |  |  |  |

### Sex differences in prevalence of OMT for IHD

Overall, the proportion of women on each recommended medication class−antiplatelet therapy, statins, β-blockers, and RAAS inhibitors was significantly lower than men. After multivariable adjustment, women had significantly lower odds of receiving antiplatelets (OR 0.71 95%CI 0.52-0.94), statins (OR 0.62, 95%CI 0.40-0.96), and RAASi (OR 0.56, 95%CI 0.38-0.84), compared with men; whereas β-blocker use did not differ significantly by sex.

Regarding combination therapies, the use of all 4 OMT medications and a combination of both antiplatelets and statins were lower among women. Even after adjusting for demographics, socio-economic factors, and comorbidities, the odds of using all 4 medications (OR 0.47, 0.28-0.79) or antiplatelets + statin therapy (OR .62, 0.42-0.89) was lower among women compared with men (**Table 2**). Differences in the use of each class or OMT combination were even more distinct among adults under 50 years. (**Table S2**)

**Table 2:** Distribution and Association of Sex and OMT Use Among Adults with Ischemic Heart Disease^a^.

| Medications | % (95%CI) |  |  |  | Adjusted Odds Ratio<sup>b</sup><br>(Women vs Men) |
| --- | --- | --- | --- | --- | --- |
|  | Women | Men | Absolute Difference | p-value |  |
| Antiplatelet | 68.0 (63.3, 72.8) | 77.7 (73.8, 81.6) | 9.7 (3.4, 16.0) | <0.01 | 0.71 (0.52, 0.94) * |
| Statin | 57.2 (52.8, 61.7) | 73.9 (70.6, 77.1) | 16.6 (11.5, 21.7) | <0.01 | 0.62 (0.40, 0.96) * |
| RAAS inhibitors | 45.6 (41.1, 50.1) | 59.0 (55.4, 62.6) | 13.4 (7.7, 19.1) | <0.01 | 0.56 (0.38, 0.84) * |
| β-blockers | 51.2 (46.7, 55.7) | 61.1 (58.0, 64.2) | 9.9 (4.3, 15.6) | <0.01 | 0.76 (0.53, 1.07) |
| At least 1 | 86.6 (83.0, 90.2) | 91.4 (89.3, 93.5) | 4.9 (0.5, 9.2) | 0.02 | 1.02 (0.50, 2.10) |
| Both antiplatelet & statin | 47.4 (43.1, 51.6) | 64.1 (59.5, 68.7) | 16.7 (10.9, 22.6) | <0.01 | 0.62 (0.42, 0.89) * |
| All 4 medications | 17.5 (13.8, 21.2) | 32.5 (28.1, 36.9) | 15.0 (8.9, 21.1) | <0.01 | 0.47 (0.28, 0.79) * |
| OMT=Optimal Medical Therapy; RAAS = Renin-angiotensin-aldosterone system inhibitors; Antiplatelet includes aspirin, clopidogrel, ticagrelor, or prasugrel<br>* = p<0.05<br><sup>a</sup> All statistics are weighted to be representative of the US civilian non-institutionalized population<br><sup>b</sup> From multivariable logistic regression models adjusted for age, race/ethnicity, educational attainment, health insurance, income level, hypertension, diabetes, obesity, hyperlipidemia, smoking status, depression, and chronic kidney disease.<br>Renin-angiotensin-aldosterone inhibitors included angiotensin receptor blockers and angiotensin converting enzyme inhibitors, β-blockers included cardio selective β-blockers, carvedilol, bisoprolol, and metoprolol |  |  |  |  |  |

### Trends in use of OMT

#### Antiplatelets

Antiplatelet use remained stable among men from 2011–2020, while women experienced a decline reaching a nadir in 2013/2014, before rebounding to 70.9% by 2017/2020. (**Figure 2**) The sex difference in antiplatelet use was largest in 2013/2014 (Absolute Difference of 17.1% 95%CI, 5.6% to 28.6%, p<0.01), but was no longer significant in 2017/2020. (**Table S1**) While similar trends were observed among adults over 50 years, there was a significant decline of 37.0% (95%CI, 4.6% to 70.4%, p<0.01) among women between 2011/2012 and 2017/2020. In 2017/2020, 58.5% (95%CI 29.0% to 87.9%, p<0.01) fewer women with IHD under 50 years used an antiplatelet. (**Figure 3**)

**Figure 2:**
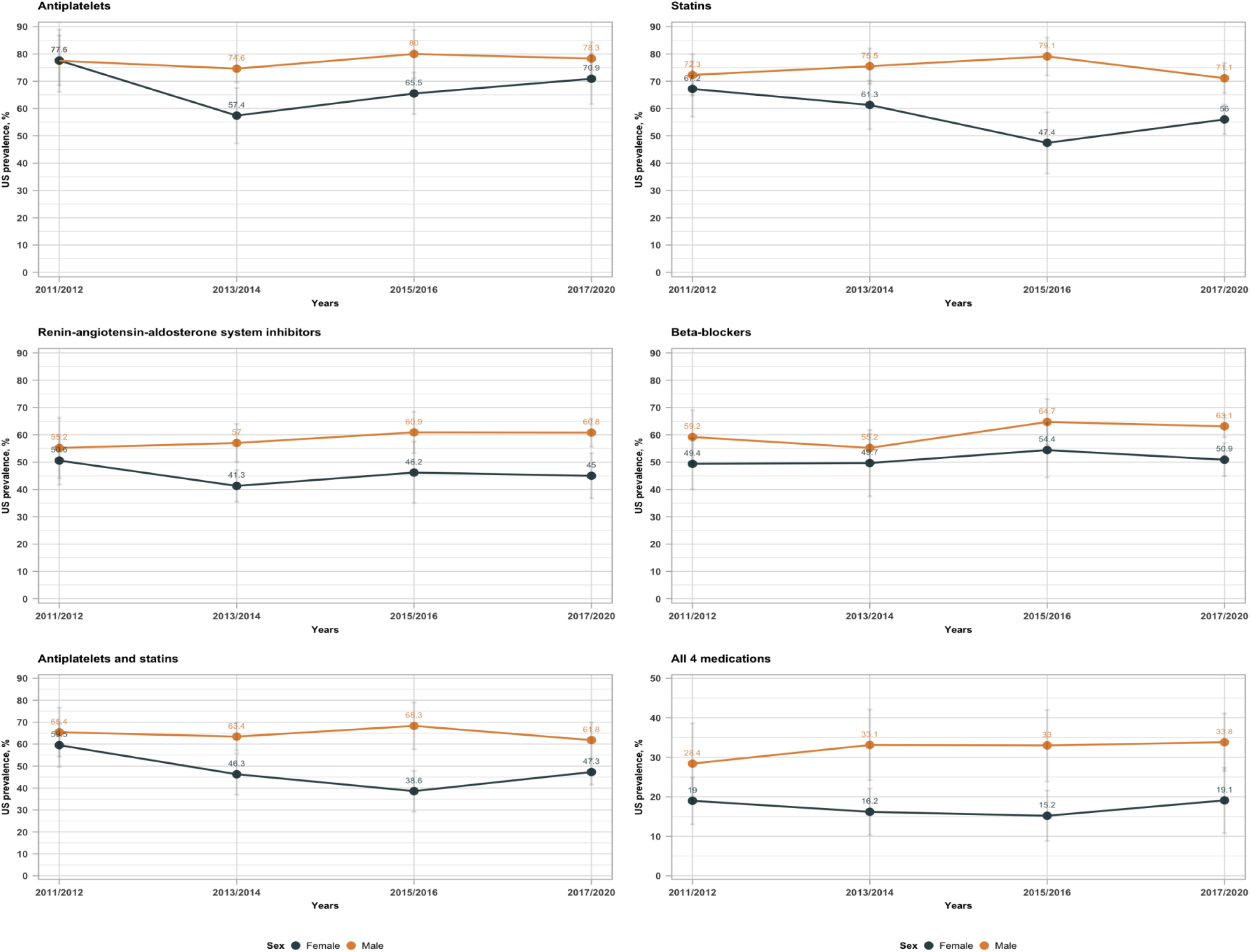
Sex differences in trends of secondary ASCVD prevention medication use across study years, 2011-2020. Antiplatelets includes self-reported and verified aspirin, clopidogrel, prasugrel, or ticagrelor use. Renin-angiotensin-aldosterone inhibitors included angiotensin receptor blockers and angiotensin converting enzyme inhibitors. β-blockers included cardio selective β-blockers: carvedilol, bisoprolol, and metoprolol.

**Figure 3:**
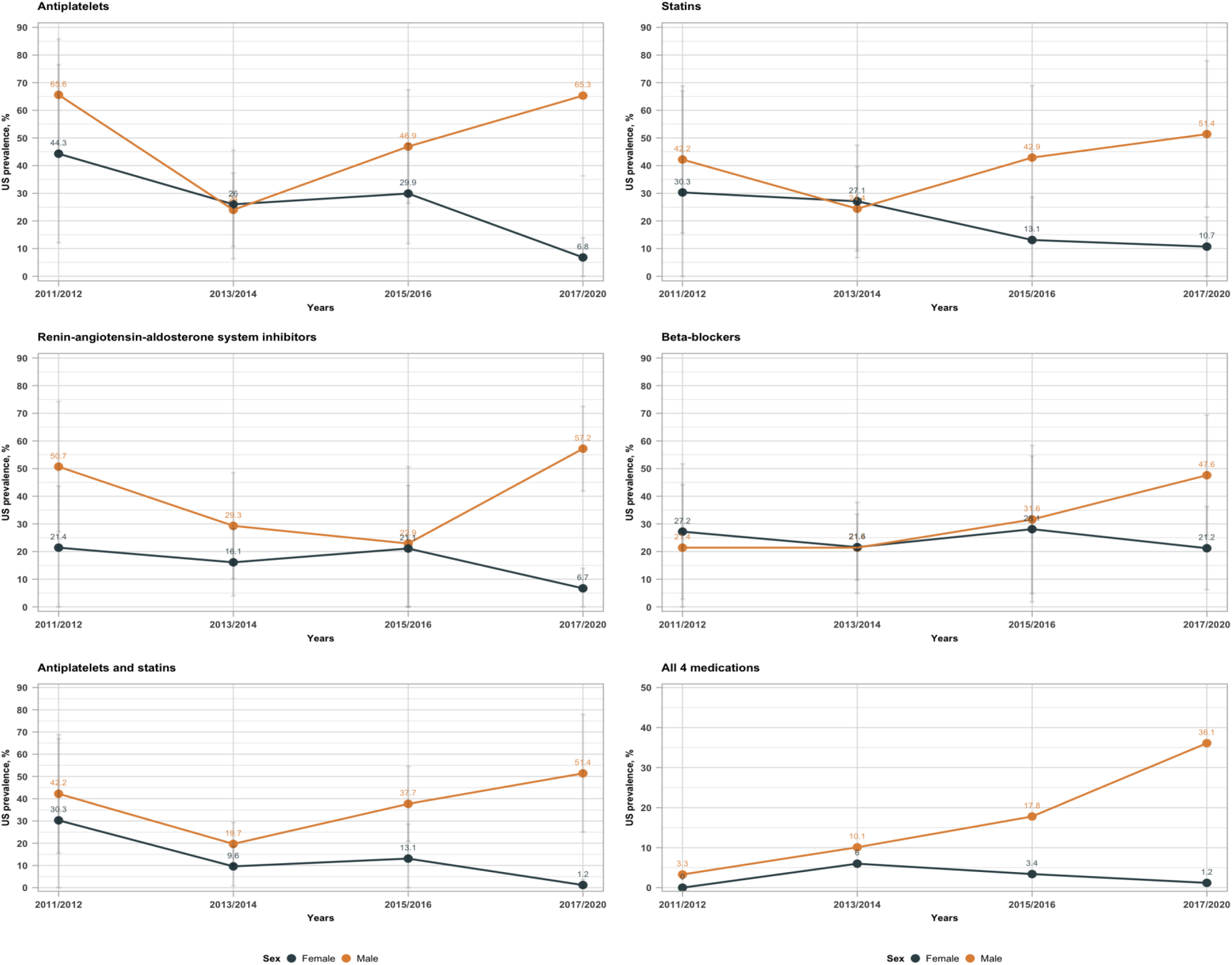
Sex differences in trends of secondary ASCVD prevention medication use across study years among adults < 50 years, 2011-2020. Antiplatelets includes self-reported and verified aspirin, clopidogrel, prasugrel, or ticagrelor use. Renin-angiotensin-aldosterone inhibitors included angiotensin receptor blockers and angiotensin converting enzyme inhibitors. β-blockers included cardio selective β-blockers: carvedilol, bisoprolol, and metoprolol.

#### Statins

In 2011/2012, 72.3% (95%CI 64.7% to 79.9%) of adult men with IHD reported using statins in the preceding month, and this proportion remained largely unchanged across the study period. Among women, 67.2% (95%CI, 57.0% to 77.3%) reported using statins in 2011/2012, and this proportion declined significantly to 56.0% (95%CI, 50.6% to 61.4%) in 2017/2020. As fewer women reported using statins, the sex difference widened, peaking at an absolute difference of 31.7% (95% CI, 20.2% to 43.1%; p < 0.01) in 2015/2016, but closed modestly in 2017/2020 (15.2% 95%CI 7.4% to 22.8%, p<0.01). (**Table S1**). Similar trends were observed in older adults. Among adults <50 years, the sex differences became wider and steady after 2013/2014, ultimately reaching 40.7% (95%CI 8.8% to 72.6%, p<0.01) in 2017/2020. (**Figure 3**).

#### RAASi

The use of RAASi was similar among both sexes in 2011/2012 (men: 55.2% 95%CI 44.1% to 66.3%, women: 50.6% 95%CI 41.7% to 59.6%), but sex differences appeared in 2013/2014 (AD 15.7% 95%CI 9.6% to 21.7%, p<0.01), owing to lower prevalence of use among women. This difference remained steady across subsequent years. (**Figure 2**) Among adults <50 years, fewer women reported using RAASi in 2017/2020, where sex differences in the use of RAASi were widest (AD 50.5 95%CI 35.9% to 65.0%, p<0.01). (**Figure 3**)

#### β-blockers

β-blocker use was generally stable over time across sexes, though a significant sex difference emerged in 2017/2020 (AD 12.2%, 95% CI, 4.3% to 20.0%) due to lower usage among women. (**Figure 2**) These trends were largely similar across age groups.

#### Both antiplatelets and statins

While the trends in the proportion of adult men and women with IHD who reported using both an antiplatelet and statin remained relatively steady across the study period; there was a significant widening of sex differences from 17.1% (95%CI 8.6% to 25.6%, p<0.01) in 2013/2014 to 29.7% (95%CI 16.1% to 43.3%, p<0.01) in 2015/2016 primarily due to a decline in the prevalence of use among women (**Figure 2**). These trends were also similar across age groups (**Figures 3 and S1**)

#### All 4 medications

Across the study cycles, a disparity between sexes in the usage of all four recommended medications was observed in 2013/2014 (AD 16.9%, 95%CI 5.5% to 28%), as well as 2017/2020, due to fewer women reporting use of all 4 medications. (**Figure 2**). In 2017/2020, only 1.2% (95% CI, 0.0% to 3.8%) of women under age 50 reported using all four recommended medications, compared to 36.0% of their male counterparts (AD 34.8%, 95% CI, 6.9% to 62.7%) (**Figure 3**).

## Discussion

In this nationally representative study of community-dwelling US adults with IHD, we found persistent and significant sex disparities in the use guideline-recommended optimal pharmacologic therapies for secondary prevention of IHD. Across the decade-long study period, women were less likely than men to receive antiplatelets, statins, and RAAS inhibitors, both as individual therapies and in combination. These differences were most pronounced among adults under 50 years of age, with younger women being particularly under-treated. Although β-blocker use did not differ significantly by sex, the underutilization of other evidence-based therapies raises substantial concerns regarding equity and outcomes in cardiovascular care.

These findings are consistent with prior reports documenting sex-based gaps in the delivery of cardiovascular care. Data from the National Ambulatory Medical Care Survey and the Veterans Health Administration have previously shown that women with atherosclerotic cardiovascular disease are less likely to be prescribed aspirin and statins than men.^5,15^ However, our study builds on this literature in several important ways. First, we observed persistent trends in sex differences across multiple drug classes and regimens. Second, we assessed longitudinal trends across a full contemporary decade, allowing evaluation of whether disparities narrowed following public awareness campaigns and policy reforms like the ACA. Finally, we incorporated age-stratified analyses, identifying younger women as a high risk group, especially vulnerable to therapeutic undertreatment.

The relatively low statin use among women—especially those under 50 years—is particularly concerning given the high burden of recurrent cardiovascular events in this demographic.^7,21^

While some clinicians may withhold statins in cases of ischemia without obstructive coronary disease—such as microvascular disease, myocardial infarction with nonobstructive coronary arteries (MINOCA), or spontaneous coronary artery dissection (SCAD)—these conditions represent only a minority of IHD cases in women.^22,23^ Moreover, many cases of MINOCA are ultimately attributable to underlying atherosclerosis, and growing evidence supports the use of statins in such contexts.^23^ Similarly, the low uptake of antiplatelet therapy including aspirin among young women is notable, especially given that women of reproductive age with a history of IHD may have dual indications for aspirin use, including preeclampsia prevention.^24^ Moreover, the low utilization of RAAS inhibitors and OMT combinations may reflect an underestimation of cardiovascular risk in women, likely driven by both provider decision-making and systemic gaps in care delivery The drivers of these disparities are likely multifactorial. Prior studies have shown that women are less likely to be offered statins, more likely to decline them, and more likely to discontinue therapy due to perceived side effects.^25^ Socioeconomic and structural barriers—including lower income, limited access to specialty care, and underinsurance—also disproportionately affect women.^26^ However, in our adjusted models, sex differences in OMT use persisted even after accounting for these access-related factors, suggesting the influence of additional unmeasured barriers. Although the Affordable Care Act (ACA) expanded coverage and preventive services for millions of Americans beginning in 2010, these gains have not eliminated longstanding disparities in cardiovascular medication use. Structural barriers in the delivery of cardiovascular care may further compound treatment inequities. Women are less likely to be referred for diagnostic coronary catheterization, and implicit bias in clinical risk assessment may lead to underestimation of cardiovascular risk and fewer evidence-based prescriptions.^27–29^ Moreover, health system structures often fail to integrate sex-specific quality measures into performance improvement efforts.

Our study highlights the need for renewed efforts to promote therapeutic equity in cardiovascular disease. Public health campaigns such as Go Red for Women have been designed to improve patient-level awareness of heart disease in women, but has not translated into universal treatment equity. Indeed, research suggests that patient-level knowledge is still lacking.^30^ Future initiatives should target both clinician education and health system accountability. Interventions may include audit-and-feedback programs with sex-specific quality metrics, embedded clinical decision support tools, and payer incentives aligned with equitable prescribing. In parallel, qualitative research is needed to better understand patient-level factors influencing medication acceptance, adherence, and long-term persistence. While our analysis focused on self-reported sex, future research should further examine the intersectionality of sex and gender to elucidate the social and biological mechanisms underlying disparities in IHD treatment and outcomes. Incorporating both dimensions may offer a more comprehensive understanding of cardiovascular risk and help tailor more equitable therapeutic approaches.

These findings are particularly timely given the current healthcare policy environment, in which proposals to scale back Medicaid funding and eligibility are under serious consideration. Medicaid is the largest payer of reproductive and preventive health services for women in the United States, and women—especially women of color and those with low incomes—are disproportionately represented among its beneficiaries.^31^ These groups are not only more likely to rely on Medicaid for access to medications and chronic disease management, but are also at higher risk of losing coverage if eligibility restrictions or budget cuts are enacted.^32^ In this context, persistent sex-based disparities in cardiovascular therapy risk becoming further entrenched unless policies are enacted to preserve and expand access to affordable care.

This study has several strengths, including the use of nationally representative data, robust statistical methodology, and evaluation of multiple pharmacologic therapies across age and time strata. However, several limitations must be acknowledged. Medication use was self-reported and subject to recall bias, though interviewers verified containers when possible. The medication list excluded less commonly used lipid-lowering agents. NHANES does not capture factors such as prescription discounts, patient refusal, or clinician rationale, which may affect observed sex differences. Although we adjusted for multiple confounders, residual confounding cannot be excluded. NHANES lacks clinical detail such as ejection fraction or angiographic data, which may inform both diagnosis and prescribing decisions. Finally, NHANES does not collect information on sex-specific risk factors, such as adverse pregnancy outcomes, which may influence cardiovascular risk assessment in women.

### Conclusions

Despite national efforts to raise awareness and broaden access to care, women with IHD— particularly younger women—remain less likely than men to receive optimal medical therapy. These disparities persist across pharmacologic classes and have not appreciably improved over time. Targeted interventions addressing structural barriers, clinical inertia, and patient-level factors are urgently needed to ensure equitable delivery of evidence-based cardiovascular care.

## Data Availability

All data produced are available online at: https://wwwn.cdc.gov/nchs/nhanes/default.aspx

## Access to data and analysis

HAA had full access to all the data in the study and takes responsibility for the integrity and the accuracy of the data analysis

## Supplemental Tables and Figures

**Figure S1:**
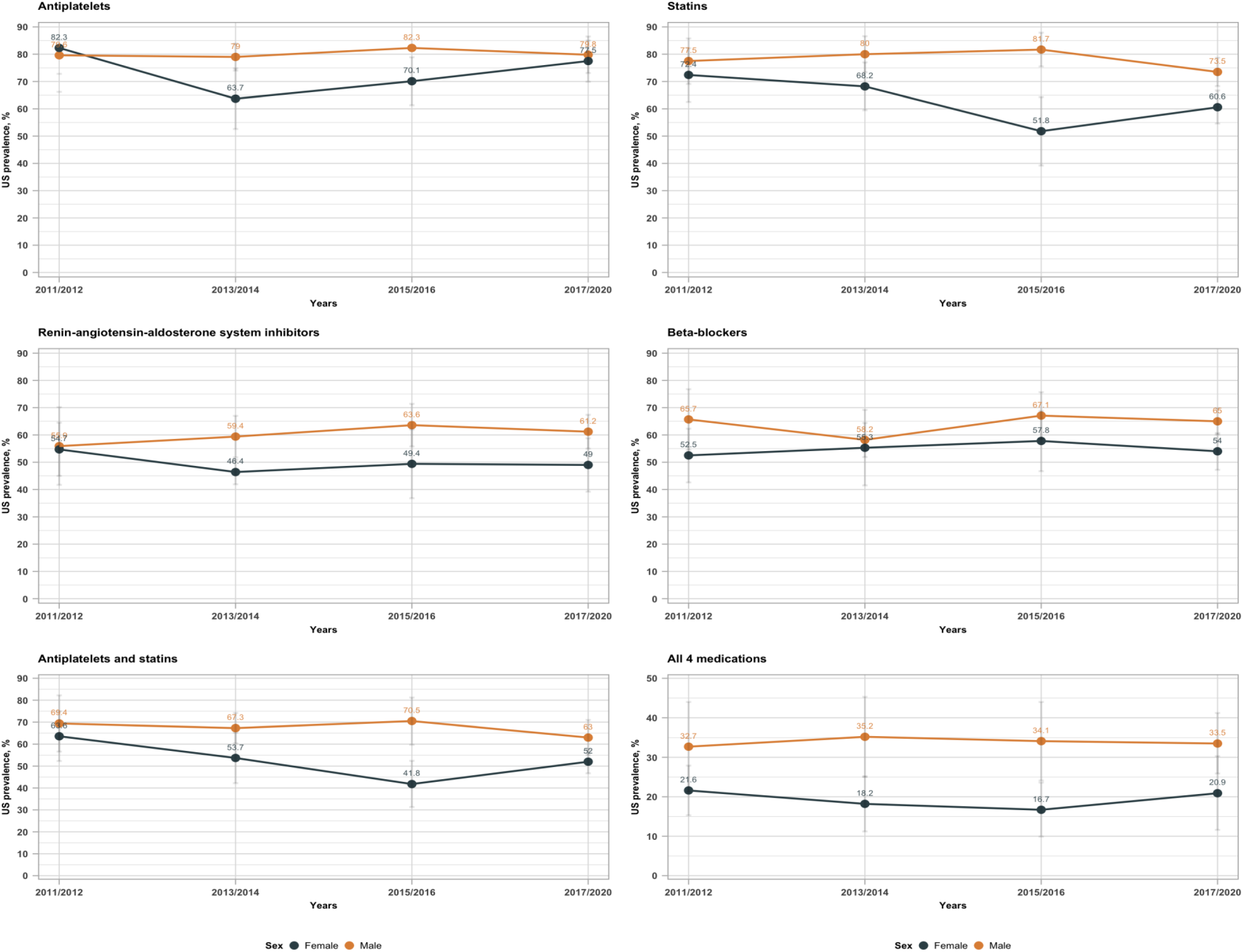
Sex differences in trends of secondary ASCVD prevention medication use across study years among adults ≥ 50 years, 2011-2020. Antiplatelets includes self-reported and verified aspirin, clopidogrel, prasugrel, or ticagrelor use. Renin-angiotensin-aldosterone inhibitors included angiotensin receptor blockers and angiotensin converting enzyme inhibitors. β-blockers included cardio selective β-blockers: carvedilol, bisoprolol, and metoprolol.

**Figure S2:**
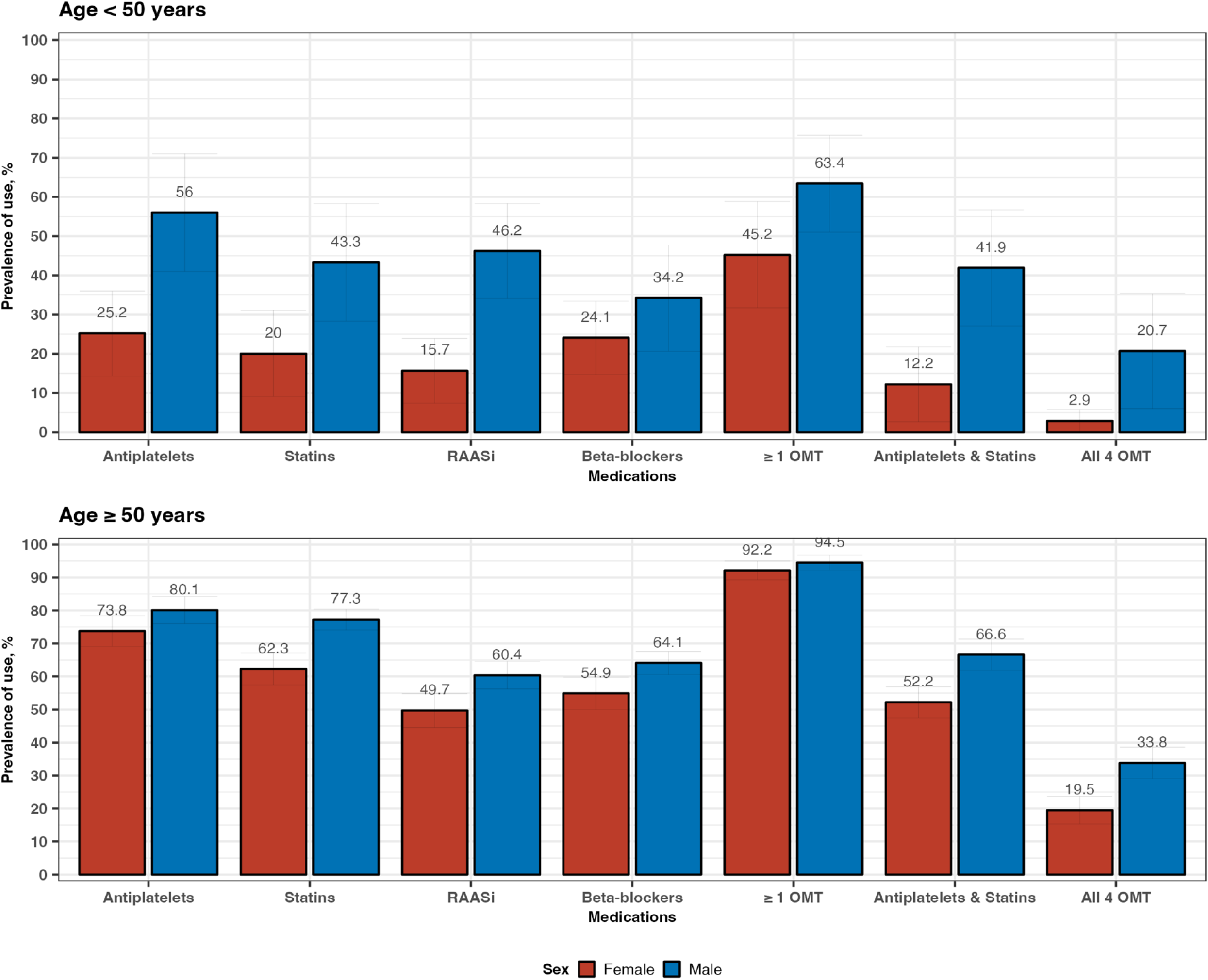
Distribution of optimal medical therapies across sex stratified by age categories-less than 50 or greater than or equal to 50. OMT implies Optimal Medical Therapy. Antiplatelets includes self-reported and verified aspirin, clopidogrel, prasugrel, or ticagrelor use. Renin-angiotensin-aldosterone inhibitors included angiotensin receptor blockers and angiotensin converting enzyme inhibitors, β-blockers included cardio selective β-blockers, carvedilol, bisoprolol, and metoprolol

**Table S1:**
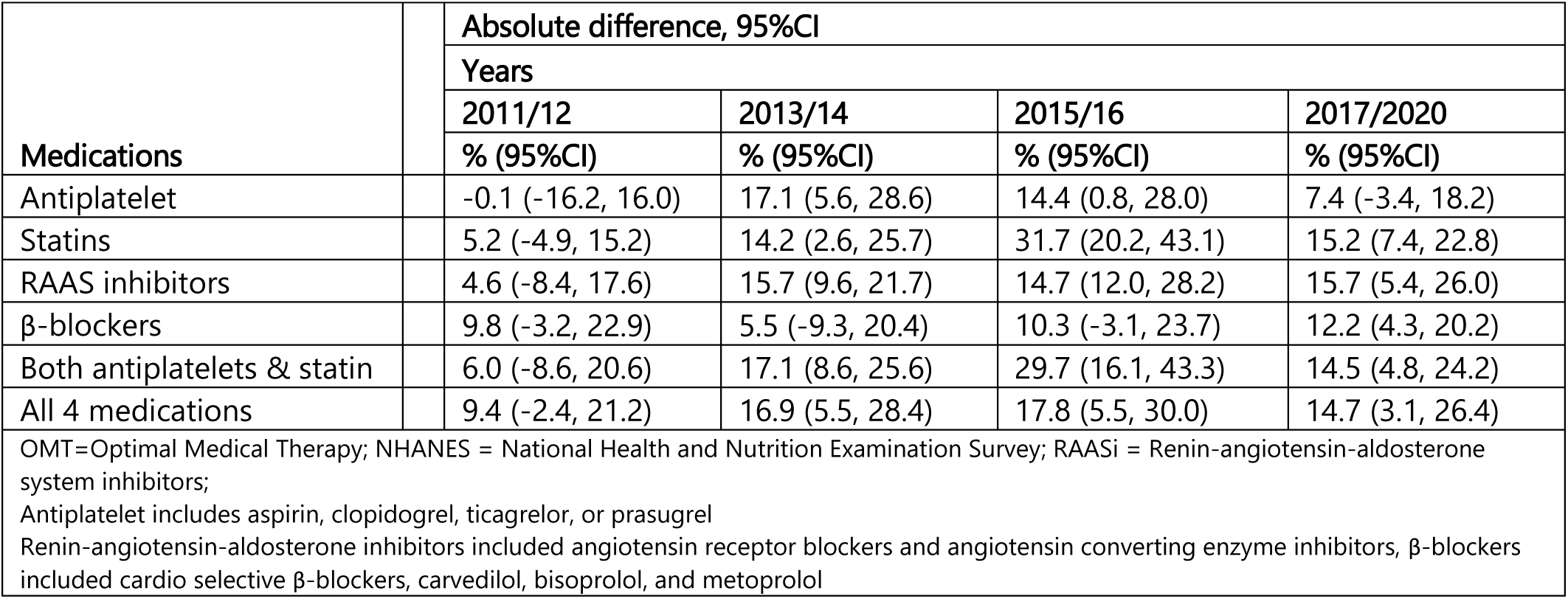
Sex difference in OMT (Men versus Women) across study years in NHANES.

| Medications | Absolute difference, 95%CI |  |  |  |
| --- | --- | --- | --- | --- |
|  | Years |  |  |  |
|  | 2011/12 | 2013/14 | 2015/16 | 2017/2020 |
|  | % (95%CI) | % (95%CI) | % (95%CI) | % (95%CI) |
| Antiplatelet | -0.1 (-16.2, 16.0) | 17.1 (5.6, 28.6) | 14.4 (0.8, 28.0) | 7.4 (-3.4, 18.2) |
| Statins | 5.2 (-4.9, 15.2) | 14.2 (2.6, 25.7) | 31.7 (20.2, 43.1) | 15.2 (7.4, 22.8) |
| RAAS inhibitors | 4.6 (-8.4, 17.6) | 15.7 (9.6, 21.7) | 14.7 (12.0, 28.2) | 15.7 (5.4, 26.0) |
| β-blockers | 9.8 (-3.2, 22.9) | 5.5 (-9.3, 20.4) | 10.3 (-3.1, 23.7) | 12.2 (4.3, 20.2) |
| Both antiplatelets & statin | 6.0 (-8.6, 20.6) | 17.1 (8.6, 25.6) | 29.7 (16.1, 43.3) | 14.5 (4.8, 24.2) |
| All 4 medications | 9.4 (-2.4, 21.2) | 16.9 (5.5, 28.4) | 17.8 (5.5, 30.0) | 14.7 (3.1, 26.4) |
| OMT=Optimal Medical Therapy; NHANES = National Health and Nutrition Examination Survey; RAASi = Renin-angiotensin-aldosterone system inhibitors;<br>Antiplatelet includes aspirin, clopidogrel, ticagrelor, or prasugrel<br>Renin-angiotensin-aldosterone inhibitors included angiotensin receptor blockers and angiotensin converting enzyme inhibitors, β-blockers included cardio selective β-blockers, carvedilol, bisoprolol, and metoprolol |  |  |  |  |

**Table S2:**
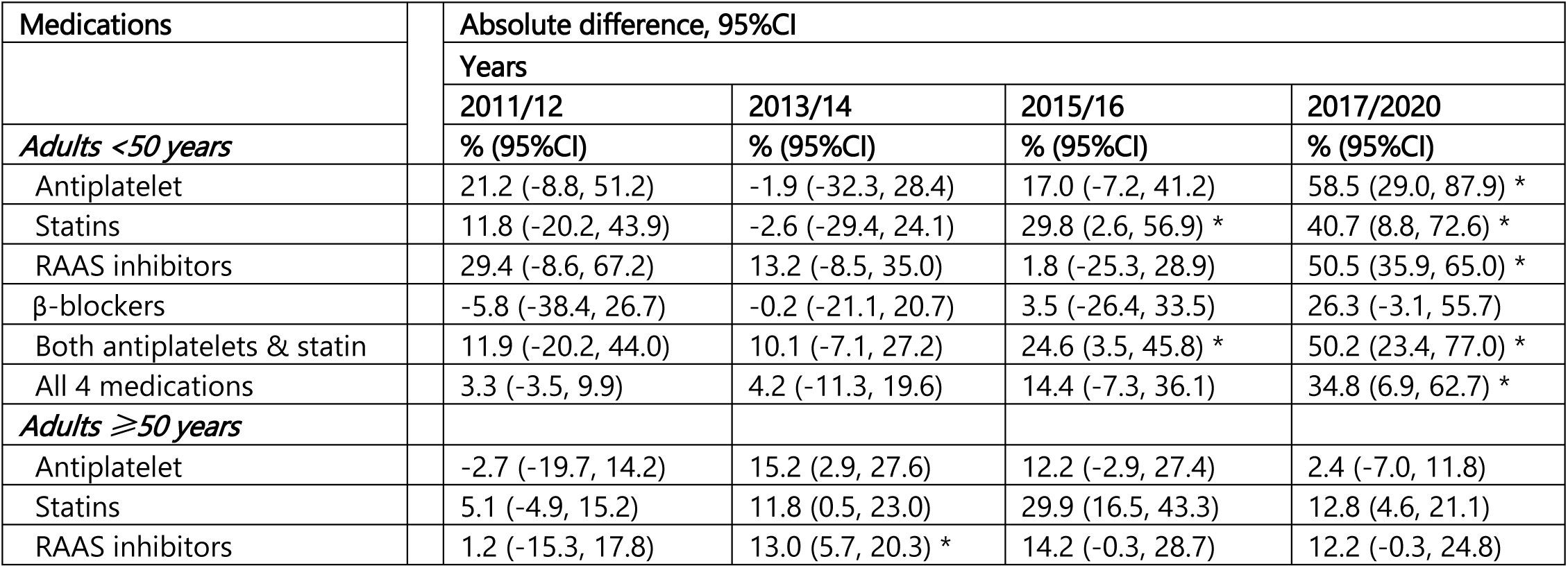
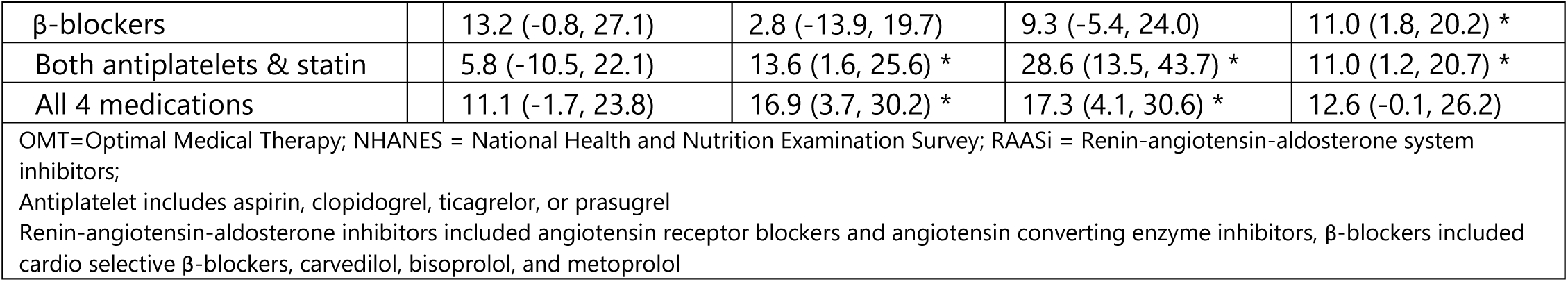
Sex difference in OMT (Men versus Women) across study years stratified by age groups in NHANES.

| Medications | Absolute difference, 95%CI |  |  |  |
| --- | --- | --- | --- | --- |
|  | Years |  |  |  |
|  | 2011/12 | 2013/14 | 2015/16 | 2017/2020 |
|  | % (95%CI) | % (95%CI) | % (95%CI) | % (95%CI) |
| **Adults <50 years** |  |  |  |  |
| Antiplatelet | 21.2 (-8.8, 51.2) | -1.9 (-32.3, 28.4) | 17.0 (-7.2, 41.2) | 58.5 (29.0, 87.9) * |
| Statins | 11.8 (-20.2, 43.9) | -2.6 (-29.4, 24.1) | 29.8 (2.6, 56.9) * | 40.7 (8.8, 72.6) * |
| RAAS inhibitors | 29.4 (-8.6, 67.2) | 13.2 (-8.5, 35.0) | 1.8 (-25.3, 28.9) | 50.5 (35.9, 65.0) * |
| β-blockers | -5.8 (-38.4, 26.7) | -0.2 (-21.1, 20.7) | 3.5 (-26.4, 33.5) | 26.3 (-3.1, 55.7) |
| Both antiplatelets & statin | 11.9 (-20.2, 44.0) | 10.1 (-7.1, 27.2) | 24.6 (3.5, 45.8) * | 50.2 (23.4, 77.0) * |
| All 4 medications | 3.3 (-3.5, 9.9) | 4.2 (-11.3, 19.6) | 14.4 (-7.3, 36.1) | 34.8 (6.9, 62.7) * |
| **Adults ≥50 years** |  |  |  |  |
| Antiplatelet | -2.7 (-19.7, 14.2) | 15.2 (2.9, 27.6) | 12.2 (-2.9, 27.4) | 2.4 (-7.0, 11.8) |
| Statins | 5.1 (-4.9, 15.2) | 11.8 (0.5, 23.0) | 29.9 (16.5, 43.3) | 12.8 (4.6, 21.1) |
| RAAS inhibitors | 1.2 (-15.3, 17.8) | 13.0 (5.7, 20.3) * | 14.2 (-0.3, 28.7) | 12.2 (-0.3, 24.8) |

|  |  |  |  |  |  |
| --- | --- | --- | --- | --- | --- |
| β-blockers |  | 13.2 (-0.8, 27.1) | 2.8 (-13.9, 19.7) | 9.3 (-5.4, 24.0) | 11.0 (1.8, 20.2) * |
| Both antiplatelets & statin |  | 5.8 (-10.5, 22.1) | 13.6 (1.6, 25.6) * | 28.6 (13.5, 43.7) * | 11.0 (1.2, 20.7) * |
| All 4 medications |  | 11.1 (-1.7, 23.8) | 16.9 (3.7, 30.2) * | 17.3 (4.1, 30.6) * | 12.6 (-0.1, 26.2) |
| OMT=Optimal Medical Therapy; NHANES = National Health and Nutrition Examination Survey; RAASi = Renin-angiotensin-aldosterone system inhibitors;<br>Antiplatelet includes aspirin, clopidogrel, ticagrelor, or prasugrel<br>Renin-angiotensin-aldosterone inhibitors included angiotensin receptor blockers and angiotensin converting enzyme inhibitors, β-blockers included cardio selective β-blockers, carvedilol, bisoprolol, and metoprolol |  |  |  |  |  |

